# How efficient are the lockdown measures taken for mitigating the Covid-19 epidemic?

**DOI:** 10.1101/2020.06.02.20120089

**Authors:** Samson Lasaulce, Vineeth Varma, Constantin Morarescu, Siying Lin

## Abstract

Various lockdown measures have been taken in different countries to mitigate the Covid-19 pandemic. But, for citizens, it is not always simple to understand how these measures have been taken. Should they have been more (or less) restrictive? Should the lockdown period have been longer (or shorter)? What would have been the benefits of starting to confine the population earlier? To provide some elements of response to these questions, we propose a simple behavior model for the government decision-making operation. Although simple and obviously improvable, the proposed model has the merit to implement in a pragmatic and insightful way the tradeoff between health and macroeconomic aspects. For a given tradeoff between the assumed cost functions for the economic and health impacts, it is then possible to determine the best lockdown starting date, the best lockdown duration, and the optimal severity levels during and after lockdown. The numerical analysis is based on a standard SEIR model and performed for the case of France but the adopted approach can be applied to any country. Our analysis, based on the proposed model, shows that for France it would have been possible to have just a quarter of the actual number of people infected (over [March 1, August 31]), while simultaneously having a Gross Domestic Product loss about 30% smaller than the one expected with the current policy

## 1 Introduction

In this work, our goal is to provide a simple model to measure the quality of the lockdown measures taken by a government to mitigate the health and macro-economic impacts of the Covid-19 pandemic. The quality is measured in terms of the tradeoff between the Gross Domestic Product (GDP) loss, the total number of infected people over a given period of time, and the maximum number of people requiring Intensive Care Units (ICU). To reach this objective, we propose a behavior model for governmental decision-making operations. Although we assume a simple measure for the quality of the lockdown measures and a simple dynamical model (namely, a classical susceptible-exposed-infected-removed (SEIR) model), the proposed approach is seen to be sufficient to constitute a first step into capturing and quantifying the tradeoff under consideration. In contrast with most studies conducted on the Covid-19 epidemic analysis where the primary goal is to refine the SEIR model (see e.g., [1, 2, 3, 4] to cite just a few) or employ the SEIR model for country regions (see e.g., [5]), our approach is to use the standard SEIR model for an entire country and choose a simple economic model to focus on the study of the tradeoff between health and economic aspects.

Recently, there have been several interesting studies on the economic impact of Covid-19 (see e.g.,[6, 7, 8, 9, 10]) but the tradeoff analysis is not chosen to be as the main focus, no model is provided for the behavior of the decision-maker, and unlike our work, the focus is not on 3-phase epidemic control policies. Concerning works on control, there have been some recent studies on how the lockdown strategies and quarantine can be planned in an optimal fashion [11, 12, 13]. A common feature among these works on optimal control and lockdown planning is that the policies considered, vary in time in a continuous manner, i.e., the lockdown policy is continuously evolving based on the infected population or just on time. However, from the perspective of a government, implementing such policies is not practical since it requires to change the measures say every day. This would lead to misunderstanding and frustration among the population. In contrast with these works, we restrict our attention to 3-phase epidemic control policies that is, the reproduction number *R*(*t*) is assumed to be constant over each of the three possible phases and we resort to an exhaustive search performed for a given number of possible levels for the reproduction number (see Fig. 1). At last, we note that the existing literature on epidemic control do not address the considered tradeoff. In fact, the closest contribution to this direction would be given by [14] where only generic discrete-time epidemics over multiple regions are considered, the particular 3-phase structure is not considered, and the key aspect of the tradeoff analysis is not developed and analyzed.

**Figure 1:**
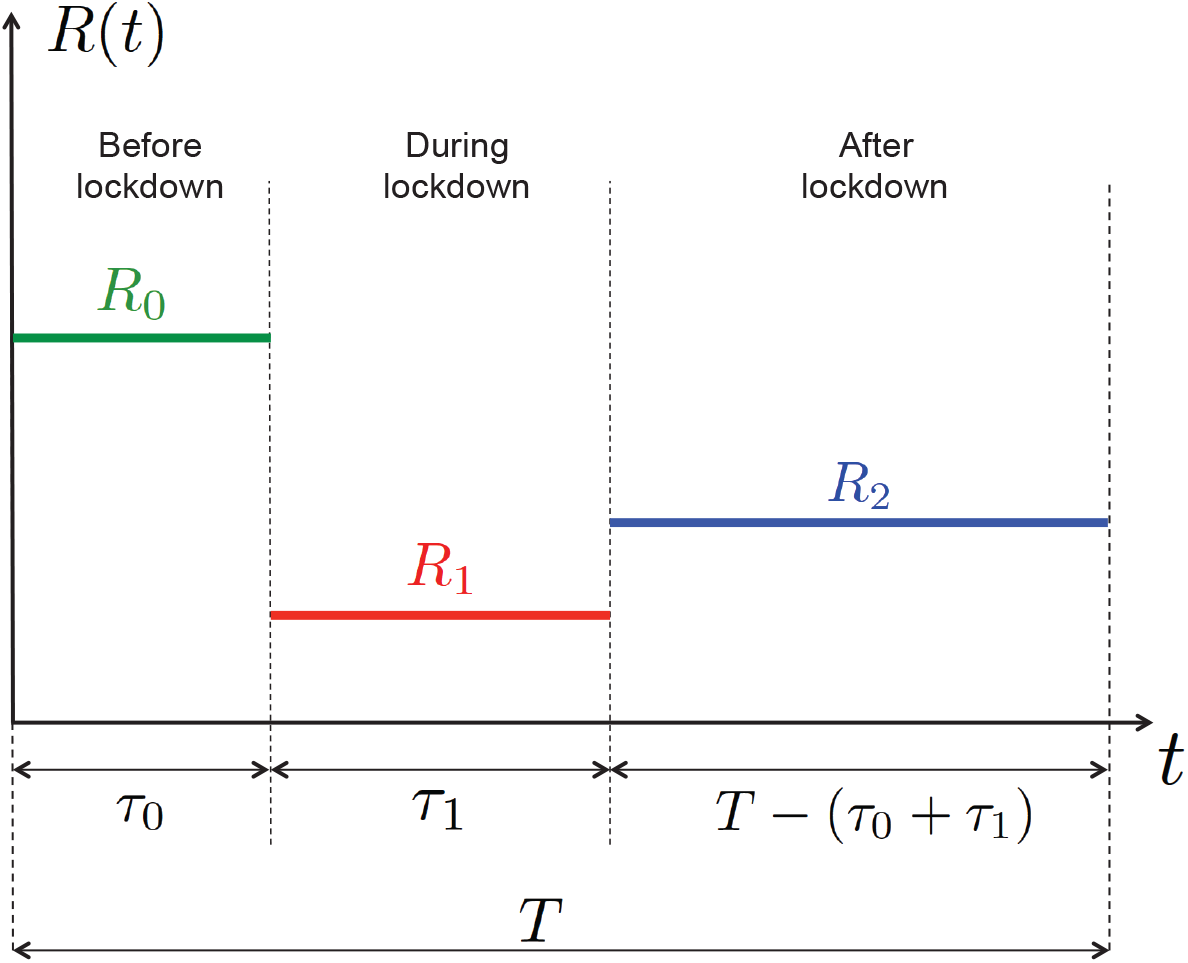
One of the goals of this work is to determine numerically, for a given tradeoff between health and economic aspects, the best 3–phase lockdown policy that is, the best values for *τ*_0_, *τ*_1_, *R*_1_, and *R*_2_ (the epidemic time horizon *T* being given and the natural reproduction number *R*_0_ being fixed).

Summarizing, compared to the existing literature on epidemics and control of epidemics, in particular, the work reported here possesses three distinguishing features:

- the focus is on 3-phase control policies and not on general continuous-time control policies;
- a simple model for capturing the tradeoff between health and economic aspect is proposed and studied;
- the numerical analysis of the tradeoff is dedicated to the Covid-19 pandemic and a case study for France.

## 2 Model

### 2.1 Epidemic model

To model the dynamics of the Covid-19 epidemic globally i.e., over an entire country, we assume a standard SEIR model. By respectively denoting *s*, *e*, *i*, and *r* the fraction of the population being susceptible to be infected by the SARS-Cov2 virus, the fraction having been exposed to it, the fraction infected, and the fraction removed (including recoveries and deceases), the epidemic is assumed to obey the following continuous-time dynamics:

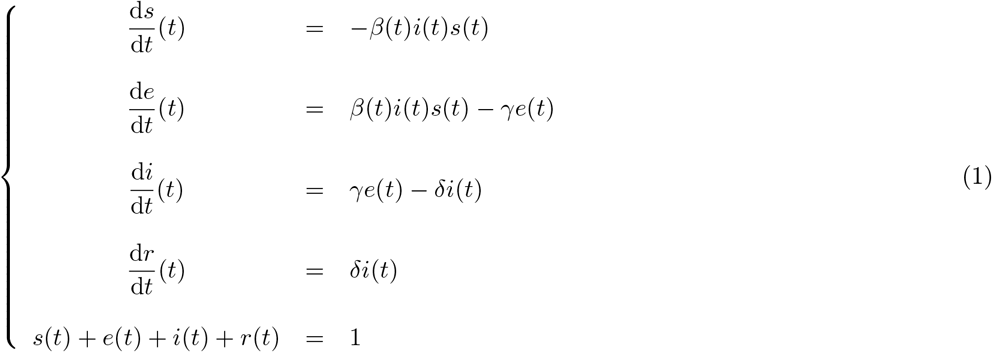

where:

- *β*(*t*), *t* ∊ ℝ, represents the **time-varying** virus transmission rate;
- *γ* denotes the rate at which the exposed subject develops the disease (this includes people presenting symptoms and asymptomatics). The period 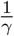 is called an incubation period;
- *δ* denotes the removal rate and 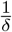 is called the average recovery period.

We assume that the the control action taken by the decision-maker (the government or possibly a more local decision-maker) has a **linear** effect on the transmission virus rate:

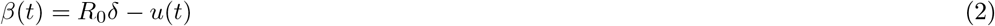

where *R*_0_ is the natural reproduction number (namely, without any control or population awareness), *u*(*t*) ∊ [0,*U*] is the control action or severity level, *U* corresponding to the most drastic or severe control action (in theory it could reach the value *R*_0_*δ* and make the reproduction number vanishing). In this work, *u*(*t*) is a piecewise-constant function. Equivalently, one can define the time-varying reproduction number:

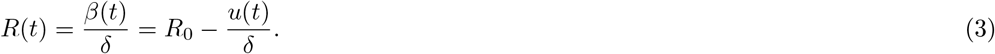

As illustrated by Fig. 1, we are solving a co-design control problem in which we are determining jointly the switching instants *τ*_0_, *τ*_1_, and the targeted reproduction numbers *R*_1_ and *R*_2_; *T* is a given period of time for the epidemic analysis.

### 2.2 Decision-maker behavior model

The proposed behavior model for the decision-maker consists in assuming that it wants to implement a given tradeoff between economic and health aspects. For the economic cost, we assume the simplest reasonable model that is, we assume that economic cost is quadratic in the control action. For the health cost, we assume that it is given by the number of infected people over the given period of time. Therefore, the proposed overall cost consists of a convex combination of these two costs. By minimizing the combined cost, one realizes the desired tradeoff. To impose the number of patients requiring intensive care, the corresponding minimization is performed under a constraint on the number of people infected at any time *t* ∊ [0, *T*]: 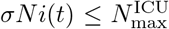, *N* being the population size, 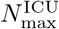 the maximum number of ICU patients, and 0 ≤ *σ ≤* 1 is the percentage of infected people requiring intensive care. In France, official records state that the maximum number of ICU patients has reached 7 148 (on April 8, 2020) but the capacity over the whole territory has been evaluated to be greater than 15 000. By denoting 0 ≤ *α* ≤ 1 the weight assigned to the macroeconomic impact of the epidemic and *K*_e_ > 0, *K*_h_ > 0, *μ >* 0 some constants (parameters) defined below, implementing the desired tradeoff amounts to fixing *α* to a given value and to find a solution to the following optimization problem (OP):

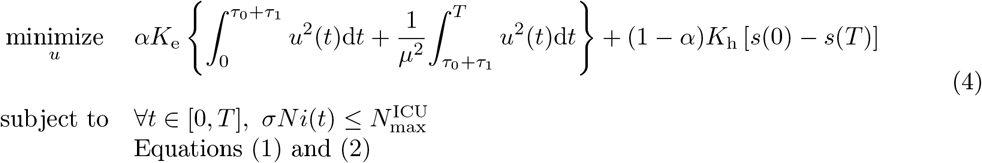

where:

- *K*_e_ > 0 and *K*_h_ > 0 are constants that weight the economic and health cost functions (they act as conversion factors allowing one to obtain appropriate units and orders of magnitude);
- *τ*_0_ and *τ*_1_ respectively represent the lockdown starting time and duration;
- the parameter *μ* ≥ 1 accounts for possible differences in terms of economic impact between the lockdown and after-lockdown phases;
- *s*(0) and *s*(*T*) are respectively the fractions of the population infected at the beginning and the end of the analysis.

We would like to make additional comments concerning the parameter *μ*. The motivation for introducing *μ* is twofold. First, after lockdown, people are more aware and act more responsible than before lockdown. This means that automatic and costless population distancing typically occurs [6, 15, 16]. Taking *μ* ≥ 1 precisely amounts to having a smaller reproduction number without any cost for the government. It also allows one to account for the fact that, after lockdown, the economic situation becomes more normal. But, here we ignore memory effects due to lockdown measures. This could be considered as a refinement of the proposed model. Perhaps, this would amount to assuming *μ* < 1, but we don’t explore this scenario in the present work.

Solving the optimization problem given by (4) is not trivial. However, since we restrict our attention to a certain class of control or lockdown policies, the problem turns out to be solvable through exhaustive search. By using the relation *u*(*t*) = *δ*[*R*_0_ − *R*(*t*)], the OP (4) can be rewritten under a more convenient form for numerical purposes:

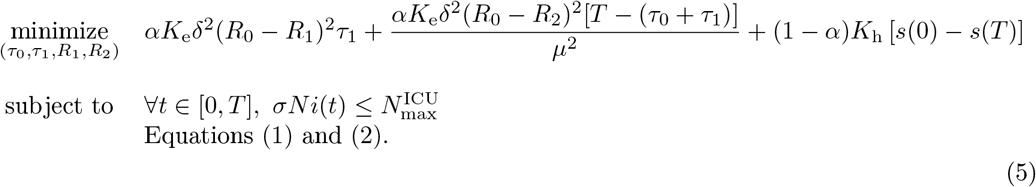

Finally, the conversion factors *K*_e_ and *K*_h_ are chosen as follows. The rationale behind the choice of *K*_e_ is that when choosing *α* =1 the GDP loss should correspond to the best estimations made by economists. The GDP loss over the lockdown period for a given country is denoted by ∆GDP, the conversion factor *K*_e_ is chosen as follows:

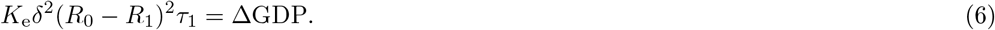

For France for example, the GDP loss during lockdown was evaluated (on April 20) to be around 120 billion € according to the OFCE [17]. At last, the constant *K*_h_ is merely chosen as *K*_h_ = *N*, that is, when *α* = 0 the cost function corresponds to the number of people infected over the considered period of time.

## 3 Numerical analysis

### 3.1 General simulation setup

To perform exhaustive search over the quadruplet of variables (*τ*_0_, *τ* _1_, *R*_1_, *R*_2_), time and amplitudes are quantized; we thus use hat notations to indicate corresponding values are quantized. Time is discretized with a step of 2.4 hours (that is, with 10 samples for each day) and a time horizon of 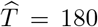 days (which approximately corresponds to 6 months that is the interval [March 1, August 31]) is assumed. The sets for the possible lockdown starting day, the lockdown duration (in days), and the reproduction numbers are as follows: 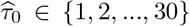, 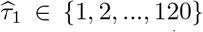, 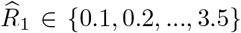, 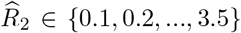. The SEIR model parameters are as follows: 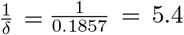, 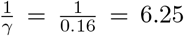; these choices are consistent with many works and in particular the studies performed for France [18] and Italy [13]. The population size is set to *N* = 66.10^6^, the maximum number of patients requiring intensive care is set to 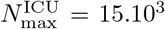, and *σ* = 1.5%. Notice that this number is only reached for very small values of *α* (for which the total number of people infected over the analysis duration would be around 9 millions). The initial number of infected people on March 1 2020 is set to *Ni*(0) = 1.33.10^5^. This number is obtained from analyzing the data provided in [19]. Indeed, the number of deceases observed at a given time is more reliable than the reported number of infected people. Therefore, by fixing the mortality rate to a given value (in [18] for instance, the mortality rate within the class of infected people is evaluated to be around 0.53% for France), one can estimate from the number of deceases at a given time the number of infected people 3 to 4 weeks before. Now concerning the economic cost for France, the GDP loss over the lockdown period is estimated by OFCE [17] to 120 billions € and we have that 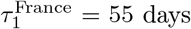 with 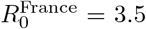 and 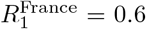. This gives *K*_e_ = 7.379.10^9^ €/day.

### 3.2 Optimal tradeoff between economic and health impacts

Choosing a value for the parameter *α* amounts to implementing a given tradeoff between the economic cost and the health cost. For *μ* = 1.41 (that is, *μ* ^2^ ~ 2), Figure 2 depicts for various values of α in the interval [10^−7^,10^−4^] the total GDP loss and number of infected people that is obtained after choosing the (quantized version of the) quadruplet (*τ*_0_, *τ*_1_, *R*_1_, *R*_2_) that minimizes the combined cost given by Equation (5). At one extreme, when α is relatively large (*α* = 10^−4^), we see that the best lockdown strategy leads to a GDP loss over the entire study period [March 1, August 31] of 125 billions € with about 9.5 millions infected, and 15 000 patients requiring intensive care. At the other extreme, when α is relatively small (*α* = 10^−7^), the GDP loss reaches values as high as 260 billion € with a total number of newly infected people over the period [March 1, August 31] as low as 48 180.

**Figure 2:**
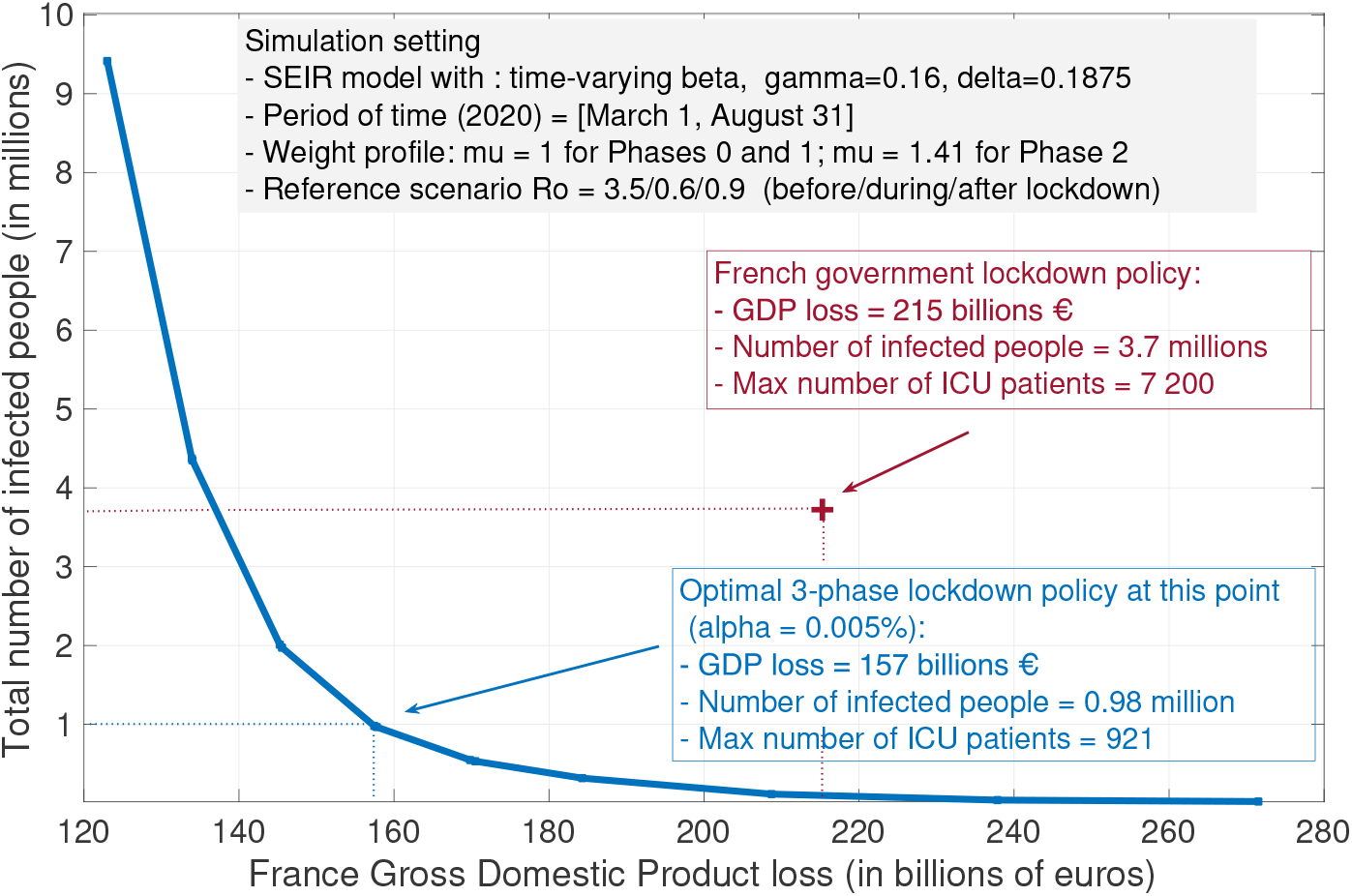
The curve represented here represents the pair best possible tradeoffs between economic cost (measured in terms of GDP loss) and health cost (measured in terms of the total number of infected people) that can be obtained (by choosing the best lockdown policy). In particular, with the assumed model, it is seen that it would have been possible to divide the number of infected people by about 4 and saving about 60 billions € in terms of GDP.

To evaluate the efficiency of the lockdown efficiency of the French government policy, we have represented the point corresponding to the estimated number of infected people and GDP loss by August 31; with our model, the GDP loss over the period of time of interest is 215 billion € and the total number of infected people is about 3.7 million. What the best tradeoff curve indicates is that there were lockdown policies that would allow the French government to have a better “performance” both in terms of GDP loss and the number of infected people. For instance, we indicate a point for which it was possible to have less than 1 million people infected (that is, about 4 times less than what is expected from the current policy) while ensuring a total GDP loss of 157 billions €(which corresponds to about 30% less than the current policy). Which type of lockdown policies should be used to have such an outcome? The next subsections provide a detailed analysis of the optimal lockdown parameters.

### 3.3 Optimal lockdown starting time

From now on, we look at the features of the optimal lockdown policy which implements a given tradeoff between economic and health costs. For *μ* =1 and *μ* = 1.41, Figure 3 provides the best day to start locking down the population, for one hundred values of α ranging from 10^-4^ to 10^-6^. When the economic cost associated with a given intensity or severity level is lower after lockdown than during it (here when *μ* = 1.41), it is always optimal to start locking down as soon as possible, which means a start on March 1 (versus March 17 in France). This result is not surprising, the sooner the better for controlling the virus expansion. Now, when economic losses are assumed to be uniform over time (*μ* = 1), there is some economic incentive to delay the lockdown, but this delay is seen to be at maximum of 3 days for the considered realistic range for *α*. Of course, our model does not capture the possible fact that people need to be psychologically prepared to follow the lockdown measures. By March 17, there were official figures about the epidemic which were sufficiently critical to make the population accept the measures whereas, starting on March 4 (the optimal starting date for *μ* =1) the situation might have not been critical enough to create full adhesion to government measures.

**Figure 3:**
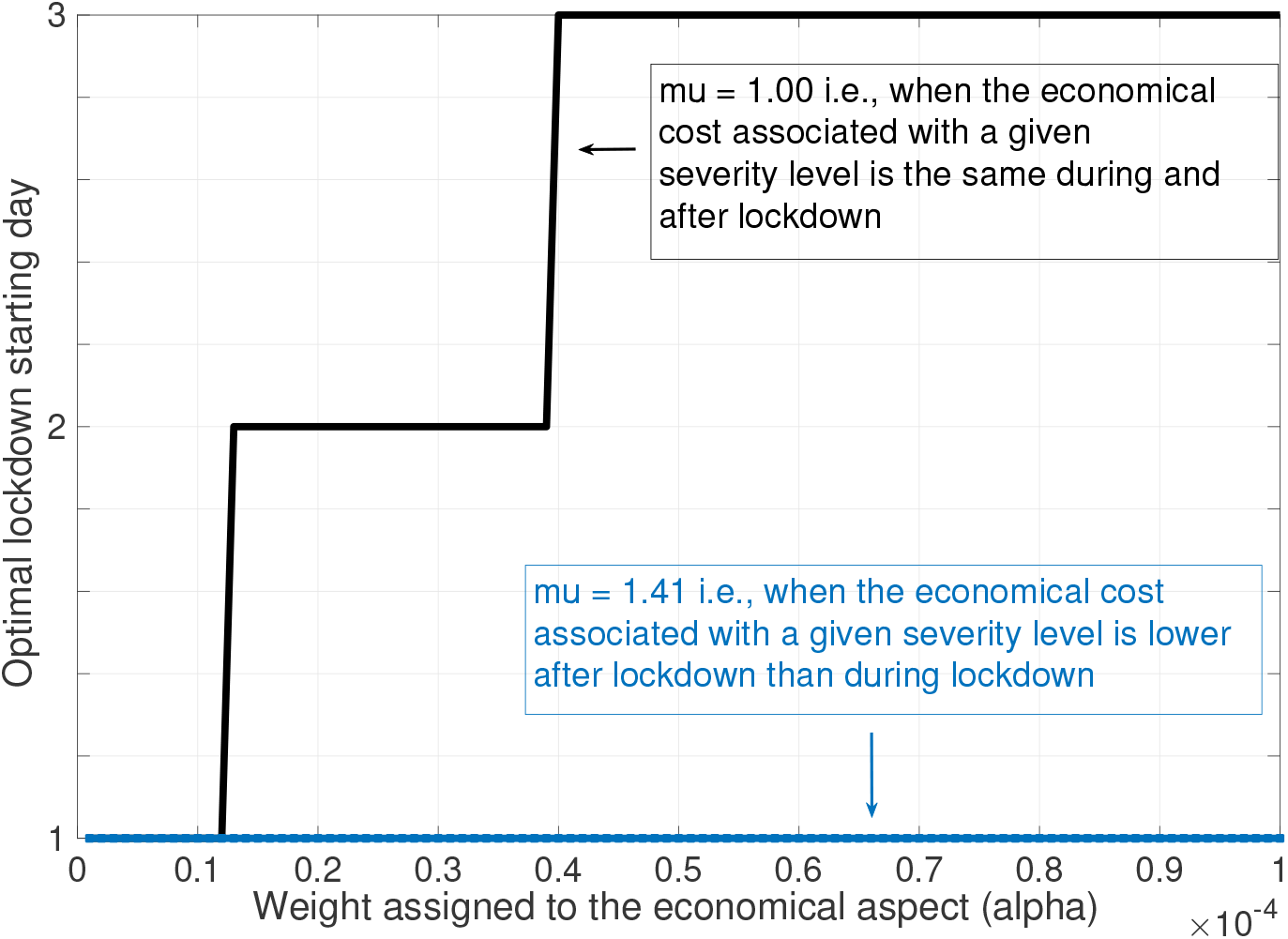
When economic losses are assumed to be uniform over time (*μ* = 1), there is some economic incentive to delay the lockdown, but this delay is seen to be at maximum 3 days. When economic losses are lower after lockdown than during it, the lockdown phase should always (up to the psychological dynamics) start as soon as possible (here on March 1).

### 3.4 Optimal lockdown duration

For *μ* =1, *μ* = 1.22, and *μ* = 1.41, Figure 4 provides the best lockdown duration (in days) for values of α ranging from 10^−4^ to 10^−6^. Very interestingly, for *μ* =1 (i.e., when economic losses are uniform over time), the optimal duration ranges from 67 days to 85 days for a large fraction of the considered interval for *α*. We see that these values are relatively close to lockdown duration in France namely 55 days. For larger values of *μ*, the optimal lockdown duration is much smaller. Therefore, if economic losses are uniform over time, the French government policy seems to be very coherent. On the other hand, if the economic impact is smaller after lockdown, our study suggests shorter lockdown durations. In fact, our results show the existence of a critical value for the tradeoff parameter *α* above which the second phase of the management of the epidemic should not be present. This means that the optimal control consists in having only one phase instead of three; such a phase should start as soon as possible and would typically last 5 – 6 months. During this phase, the population would have to be in an intermediate state in which the reproduction number is sufficiently low (about 2 as seen further). We see that the policy would be very similar to what is done for the flu with the difference that the population is informed, pro-active, and monitored. The underlying assumption here is that the epidemic would then naturally vanish (e.g., because of seasonal effects). Note that all of these observations hold under an optimal choice for the lockdown starting time (as explained in the previous subsection). If the lockdown phase starts too late, this approach can no longer be adopted.

**Figure 4:**
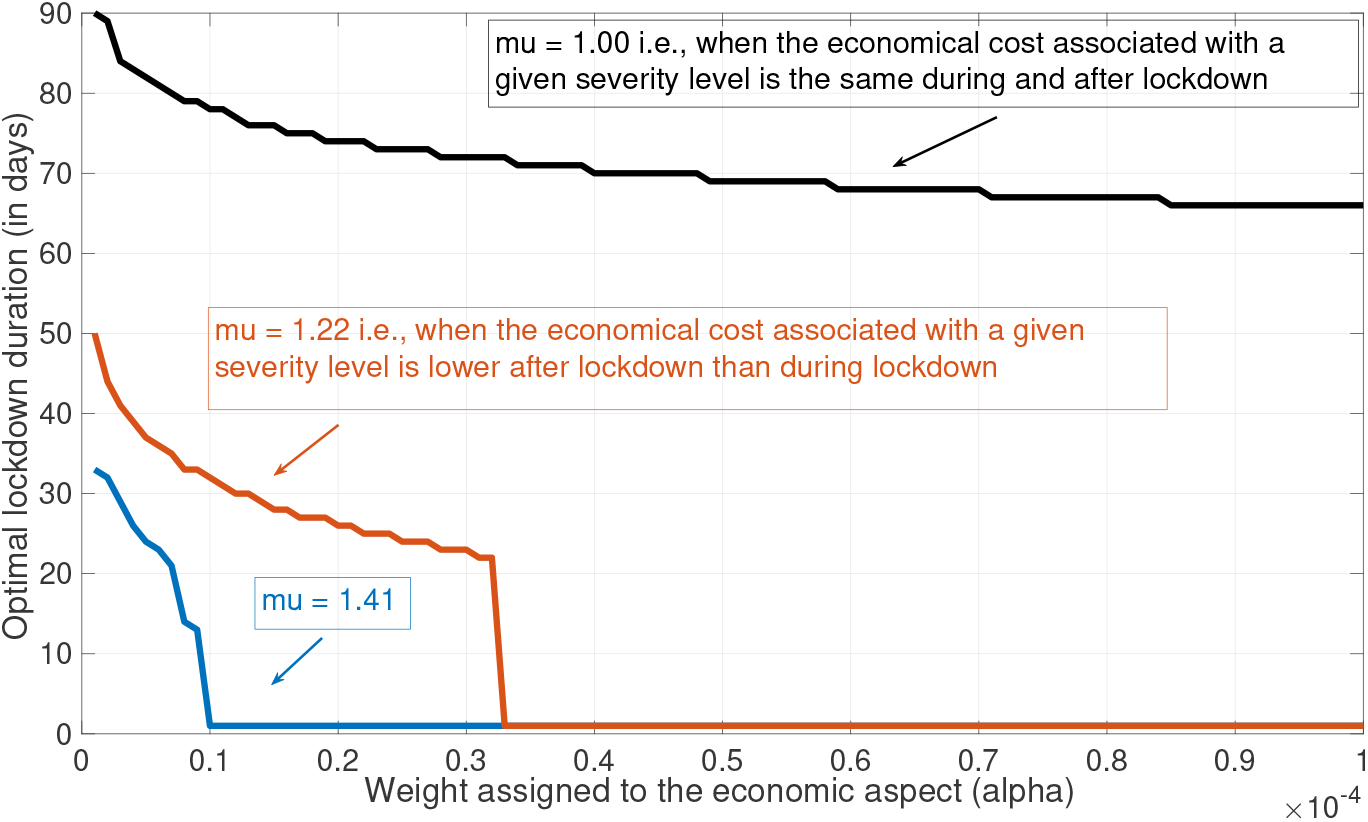
When economic losses are assumed to be uniform over time (*μ* = 1), it is optimal to have a long lockdown (typically between 60 – 80 days) whatever the tradeoff desired. But, if the economic losses of the third phase are less than during lockdown, the lockdown phase should be shorter or even absent.

### 3.5 Optimal severity levels

Figure 5 shows, for the French government policy, the evolution of the number of infected people (in millions) and the associated 3-phase reproduction number profile. Whereas, the values of the reproduction number for the first two phases come from past and quite accurate evaluations (see e.g.,[20]), the value *R*_0_ = 0.9 corresponds to the assumption that the population will do just what needs to be done to avoid a second wave. For *μ* =1 and *α* = 5.10^−5^, Figure 6 shows the optimal 3-phase profile for the reproduction number and the corresponding dynamics for the Covid-19 epidemic. Clearly, starting locking down really is one feature of this optimal profile. Then the severity level of the lockdown phase should be as high as possible, here *R*_1_ =0.1. In France, such a value might have been obtained by using more face masks, digital tracing, imposing a more severe policy for food supply. Then would follow a third phase during which the reproduction number should be around 2.1, which is perfectly achievable with natural distancing and appropriate equipment. Note that would mean the presence of a second wave, but again, this wave could be damped by phenomena like seasonal effects (e.g., effects of ultra-violets, higher dryness levels, and more uniform population density). Without any damping effect, note that the height of this wave is still 4 times lower than the one observed for Figure 5. At last, notice the presence of a significant part of the curve for which the number of infected people seems to be zero. In fact, it is not strictly zero. The minimum is naturally reached at the end of the second phase and it can be checked that the number of infected people is 48. Then, because the reproduction number is 2.1 and not 3.5 the epidemic takes some time to re-expand. In fact, the apparent second wave could thus also be damped by using appropriate detection and quarantine measures.

**Figure 5:**
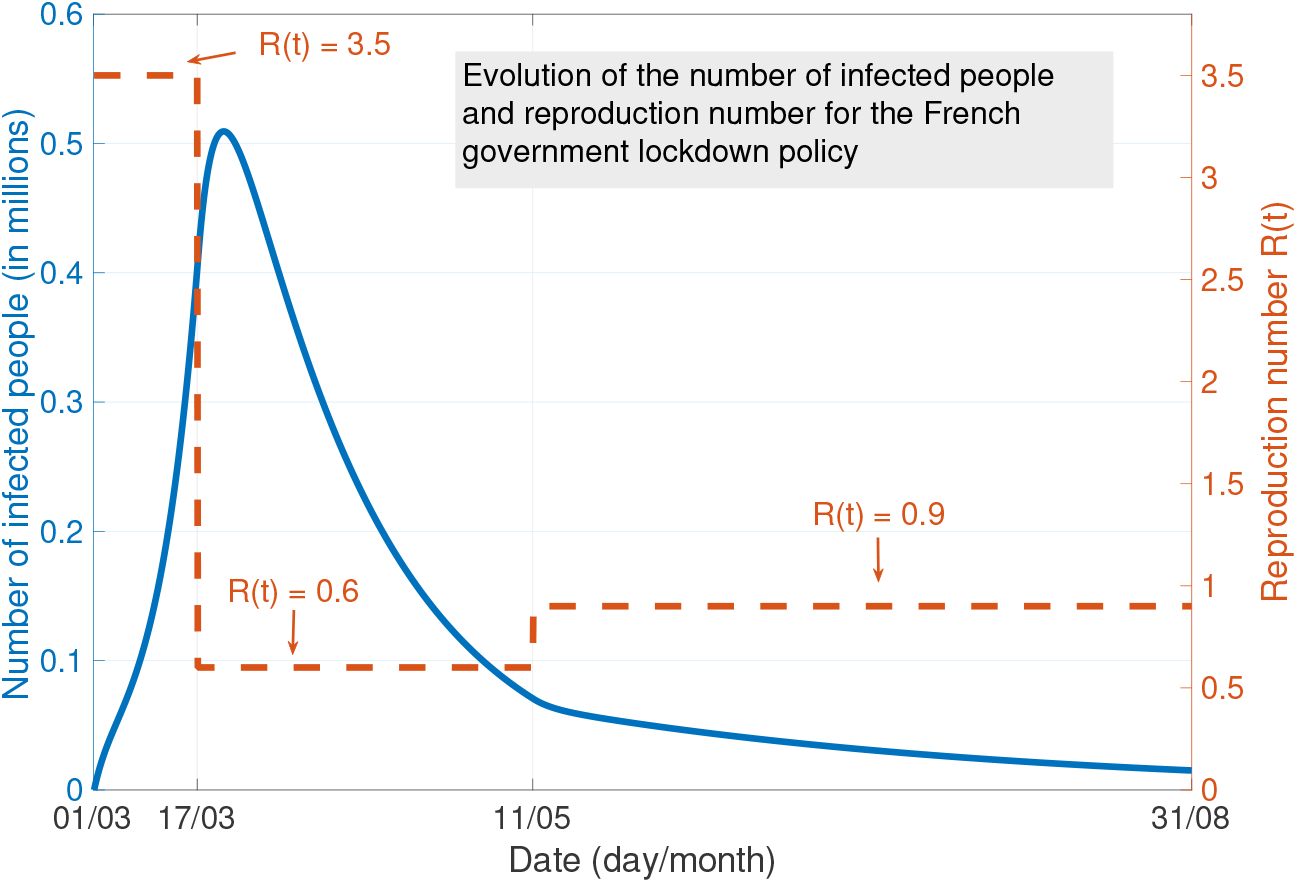
Evolution of the number of infected people and assumed reproduction number profile for the French government policy.

**Figure 6:**
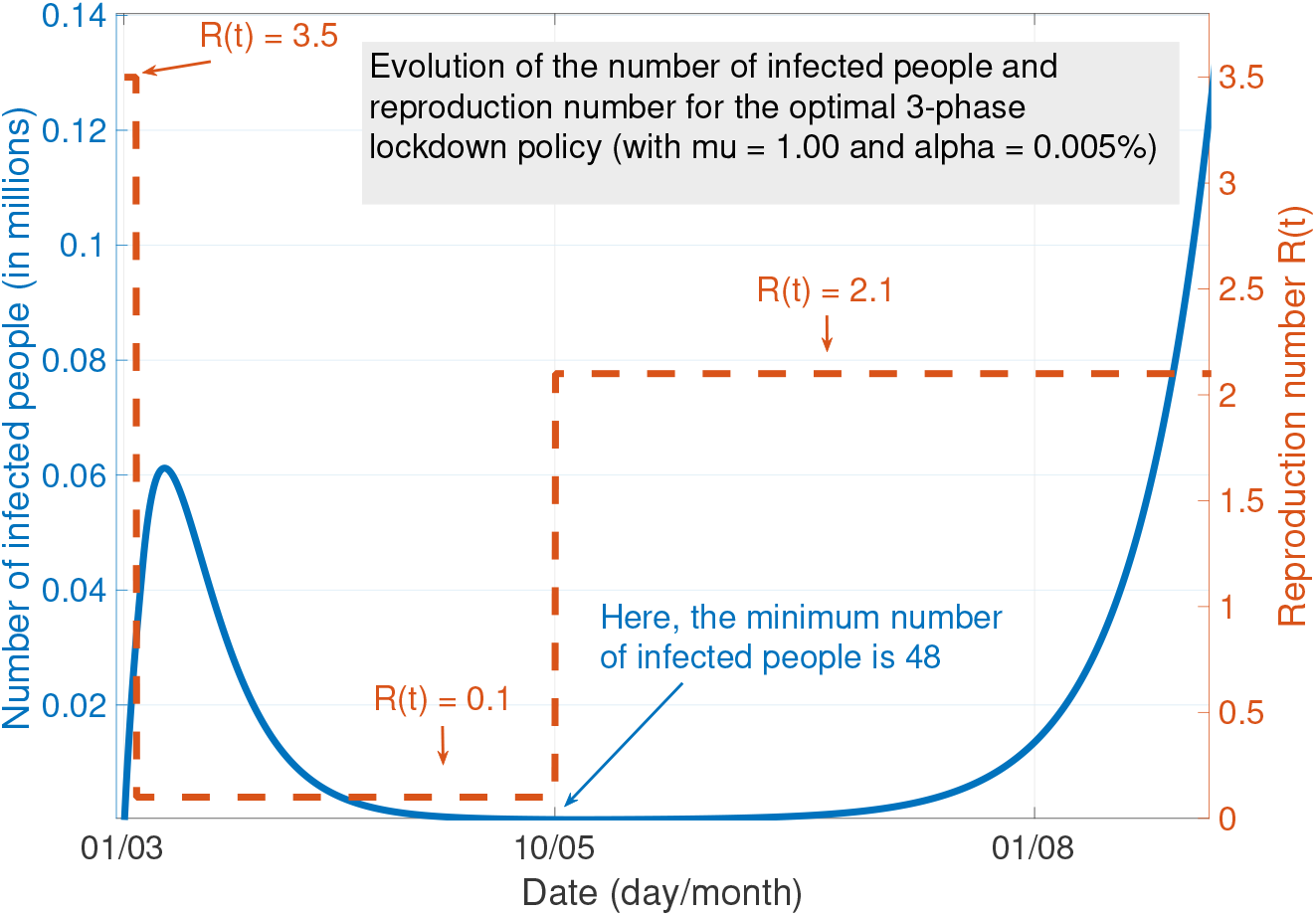
Optimal lockdown policy and corresponding dynamics for the Covid-19 epidemic.

## 4 Conclusion

In this work, we propose to model the behavior of a government as far as the lockdown policy is concerned. The proposed model, despite of its simplicity, has the merit to capture the fundamental tradeoff between economic and health aspects. By modeling the Covid-19 epidemic with a standard SEIR model, we derive the optimal parameters of the 3–phase lockdown policy. The conclusions in terms of optimal policies depend in part on whether economic losses can be considered uniform over time or are less after lockdown during it. Here are some key observations the proposed model allowed us to make:

- The lockdown phase should generally start as soon as possible. Even when economic losses are uniform over time and by putting relatively higher importance on the economic aspects, the lockdown should not be delayed by more than 3 days from the date at which the epidemic is identified (here on March 1). Here, the psychological dynamics of the population might be taken into account to refine this conclusion.
- The optimal lockdown duration found with the proposed model ranges from 60 days to 80 for a large range for the possible tradeoffs, which is close to the lockdown duration in France (55 days). But this result is obtained when economic losses are uniform over time. When economic losses are lesser after lockdown, the proposed suggest shorter lockdown duration (more in the range 20 – 40 days, and even no lockdown at all if the economic aspect becomes more important. Then, to refine this conclusion, the proposed model could be extended by accounting for the economic cost associated with the health care system.
- Concerning the optimal severity level during the lockdown, quite unsurprisingly, it should be as high as possible (*R*_1_ = 0.1 with the proposed model). Such as value would be attainable with more severe measures (e.g., by using a food supply system closer to that used in China [21, 22, 23]) and using more equipment (face masks and digital tracing in particular). The optimal value found for the phase after lockdown (called *R*_2_) is typically around 2, leading to a second wave. However, the height of the second wave is reasonable compared to what has been observed in France and since it occurs in summer, it could be damped to some extent by digital awareness control campaigns and various seasonal effects.

At last, note that we have considered an optimistic scenario for which the duration of interest for the epidemic is 6 months but, results might differ for longer periods of time.

## Data Availability

NA

## Notes

### Competing Interest Statement

The authors have declared no competing interest.

## References

[1] L. Peng, W. Yang, D. Zhang, C. Zhuge, and L. Hong. Epidemic analysis of COVID-19 in China by dynamical modeling. arXiv preprint arXiv:2002.06563, 2020.

[2] P. Lenka and V. Hajnova. SEIAR model with asymptomatic cohort and consequences to efficiency of quarantine government measures in COVID-19 epidemic. arXiv preprint arXiv:2004.02601, 2020.

[3] A. Victor. Estimation of the probability of reinfection with COVID-19 coronavirus by the SEIRUS model. *Available at SSRN 3571765*, 2020.

[4] G. Giordano, F. Blanchini, R. Bruno, P. Colaneri, A. Di Filippo, A. Di Matteo, and M. Colaneri. A SIDARTHE model of COVID-19 epidemic in Italy. arXiv preprint arXiv:2003.09861, 2020.

[5] L. Di Domenico, G. Pullano, P. Coletti, N. Hens, and V. Colizza. Expected impact of school closure and telework to mitigate COVID-19 epidemic in France. 2020.

[6] A. Atkeson. What will be the economic impact of COVID-19 in the US? rough estimates of disease scenarios. Technical report, National Bureau of Economic Research, 2020.

[7] Martin S Eichenbaum, Sergio Rebelo, and Mathias Trabandt. The macroeconomics of epidemics. Technical report, National Bureau of Economic Research, 2020.

[8] R. Baldwin and B. Weder di Mauro. Economics in the time of COVID-19. A VoxEU. org Book, Centre for Economic Policy Research, 2020.

[9] N. Fernandes. Economic effects of coronavirus outbreak (COVID-19) on the world economy. *Available at SSRN 3557504*, 2020.

[10] W. McKibbin and R. Fernando. The global macroeconomic impacts of COVID-19: Seven scenarios. Technical report, Centre for Applied Macroeconomics Analysis, 2020.

[11] F. E. Alvarez, D. Argente, and F. Lippi. A simple planning problem for covid-19 lockdown. Technical report, National Bureau of Economic Research, 2020.

[12] Harout Boujakjian. Modeling the spread of Ebola with SEIR and optimal control. SIAM Undergraduate Research Online, 9:299–310, 2016.

[13] F. Casella. Can the COVID-19 epidemic be managed on the basis of daily data? arXiv preprint arXiv:2003.06967, 2020.

[14] O. Zakary, M. Rachik, and I. Elmouki. On the analysis of a multi-regions discrete SIR epidemic model: an optimal control approach. International Journal of Dynamics and Control, 2017.

[15] M. Greenstone and V. Nigam. Does social distancing matter? Becker Friedman Institute for Economics Working Paper, 2020.

[16] M. S. Andersen. Early evidence on social distancing in response to covid-19 in the united states. *Available at SSRN 3569368*, 2020.

[17] Observatoire français des conjonctures économiques (OFCE). Évaluation au 20 avril 2020 de l’impact économique de la pandémie de COVID-19 et des mesures de confinement en France. OFCE policy brief, 2020.

[18] H. Salje, C. Kiem, N. Lefrancq, N. Courtejoie, P. Bosetti, J. Paireau, A. Andronico, N. Hoze, J. Richet, C.-L. Dubost, and et al. Estimating the burden of sars-cov-2 in france. Science Mag, 2020.

[19] Open Stats Coronavirus. Covid-19 statistiques/France. Technical report, https://www.coronavirus-statistiques.com/stats-globale/covid-19-par-pays-nombre-de-cas/, 2020.

[20] J.-F. Delfraissy, L. Atlani-Duault, D. Benamouzig, L. Bouadma, J.-L. Casanova, […], D. Malvy, and Y. Yazdanpanah.

[21] Z. Wu and J. M. Mcgoogan. Characteristics of and important lessons from the coronavirus disease 2019 (COVID-19) outbreak in China: summary of a report of 72 314 cases from the Chinese center for disease control and prevention. Jama, 2020.

[22] H. Lau, V. Khosrawipour, P. Kocbach, A. Mikolajczyk, J. Schubert, J. Bania, and T. Khosrawipour. The positive impact of lockdown in wuhan on containing the COVID-19 outbreak in China. Journal of travel medicine, 2020.

[23] H. Fang, L. Wang, and Y. Yang. Human mobility restrictions and the spread of the novel coronavirus (2019-ncov) in China. National Bureau of Economic Research, 2020.

